# Positive end expiratory pressure in invasive and non-invasive ventilation of COVID-19 acute respiratory distress syndrome: computational modeling illuminates the data

**DOI:** 10.1101/2021.11.15.21266157

**Authors:** Liam Weaver, Declan G. Bates, Luigi Camporota

## Abstract

Positive end expiratory pressure (PEEP) is routinely used as part of lung protective ventilation strategies in the treatment of acute respiratory distress syndrome (ARDS). In the case of ARDS arising due to COVID-19 (CARDS), there is some debate as to whether the atypical pathophysiological characteristics of the disease which lead to hypoxaemia could warrant a modified approach to ventilator management, particularly with regards to PEEP settings. Here we review the available evidence for the existence of a unique underlying lung pathophysiology in CARDS, and for the suitability of standard approaches to setting PEEP, in both the invasive and non-invasive ventilation settings. We show how detailed computational models informed by this evidence can shed light on the available data, and help to interpret recent results in the literature.

## Introduction

Coronavirus-19 disease (COVID-19) pneumonia has many clinical characteristics compatible with the definition of acute respiratory distress syndrome (ARDS), with bilateral lung infiltrates on chest radiology, an oxygenation defect with PaO_2_/FiO_2_ ratio < 300 mmHg and increased dead space ventilation. Positive end expiratory pressure (PEEP) is routinely used as part of lung protective ventilation strategies in the treatment of acute respiratory distress syndrome (ARDS), [1,2]. In the case of ARDS arising due to COVID-19 (CARDS), there is some debate as to whether atypical pathophysiological characteristics of the disease (i.e. moderate-severe hypoxaemia despite preserved lung volumes at presentation, and minimal parenchymal consolidation) could warrant a different approach to ventilator management, particularly with regards to PEEP settings [3,4]. Here we review the available evidence for the existence of a unique underlying lung pathophysiology in CARDS, and discuss the implications for standard approaches to setting PEEP, in both the invasive and non-invasive ventilation settings. We show how detailed computational models informed by this evidence can shed light on the available data, and help to explain recent results in the literature.

### Evidence for a unique pathophysiology underlying CARDS

There is by now abundant evidence to suggest that several of the clinical features of CARDS, particularly in its early stages, are unique, or at the very least atypical, when compared to ARDS from standard etiologies. Early stage CARDS patients typically present with focal subpleural and peri-bronchovascular ground glass opacities, very limited amounts of alveolar collapse and atelectasis, low extravascular lung water (EVLW) accumulation, and relatively well preserved compliance, features which cannot fully explain the associated large shunt fraction and severe hypoxemia [5,6]. Typically, in patients with ARDS, hypoxemia is proportional to the quantity of anatomical shunt (i.e., the fraction of non-aerated lung tissue mass in relation to the total tissue mass). The small proportion of consolidated lung tissue seems to distinguish patients with early CARDS from those with ARDS exhibiting similar PaO_2_/FiO_2_ ratio. A complete understanding of the pathophysiological mechanisms underlying CARDS is still to emerge, however the etiology of the associated early respiratory failure and disproportionate hypoxemia is likely to be due to multiple factors affecting the distribution of pulmonary perfusion in relation to areas of the lung that are more or less aerated. These factors include:

- Pulmonary vasculopathy with loss of adaptive hypoxic pulmonary vasoconstriction and dysregulated pulmonary perfusion [7,8]. Although the pivotal role ACE2 receptors play in SARS-CoV-2 transmission is well defined, their expression within the pulmonary endothelium and role in dysregulated pulmonary perfusion is now becoming apparent [7]. The carboxypeptidase ACE2 counteracts the renin-angiotensin-aldosterone system through conversion of angiotensin-I and -II to angiotensin-(1–9) and -(1–7), respectively, these then promote localised vasodilation and attenuation of the immune response [9]. The initial downregulation of ACE2 results in angiotensin-II accumulation with resulting chemotactic effects and accelerated lymphocyte recruitment [7,10]. The resulting pulmonary vascular inflammation results in an ACE1 ‘shedding’ phenomenon where endothelial surface-bound ACE1 is released into the interstitium and ultimately results in sub-physiologic angiotensin-II concentrations [10]. Low angiotensin-II concentrations in this phase lead to vasodilation and worsened capillary leak. The alterations in pulmonary perfusion determine large ventilation/perfusion inequalities with greater presence of lung compartments with low ventilation/perfusion ratio. Some of the perfusion abnormalities are “functional’ consequent to a loss in hypoxic vasocontriction and inflammatory vasoplegia leading to hyper-perfusion of poorly ventilated lung tissue (increase in venous admixture); while others are more “structural” anatomical changes caused by vascular enlargement or neovascularisation within the poorly ventilated lung parenchyma. These regions with very low ventilation/perfusion ratio can become more numerous in the presence of high cardiac output (e.g., fever and hyperinflammation) leading to hyperperfusion of poorly ventilated alveolar units.
- The high incidence of pulmonary microvascular and macrovascular thrombosis offers insight into the high compliance, increased dead space, D-dimer elevation, and right ventricular dysfunction frequently observed in COVID-19 and documented in post-mortem findings and histology [7,11,12,13,14].
- The neurotropic potential of SARS-CoV-2 with altered central control of breathing mediated by pontine pneumotaxic centre dysfunction and the relatively normal lung volume which results in increased tidal volumes relative to ventilatory frequency. Although low pulmonary elastance partially explains the deceptively effortless work of breathing [15], infiltration of SARS-CoV-2 into the cerebrospinal fluid (CSF), carotid body sensing and impaired brainstem autoregulation may also contribute [16,17].
- Increased basal metabolic rate resulting in higher tissue oxygen extraction, lower mixed venous oxygen content, and increased venous admixture [18].
- Increased intrapulmonary shunt fraction with cardiac output elevation [19,20]. Although high peripheral oxygen extraction partially explains the increased venous admixture observed in catabolic states, there may also be alterations in regional pulmonary blood flow distinct from this related to increased cardiac output [18,19,20].

In spite of the well documented pathophysiological features described above, many of which are unique to CARDS, there has been much controversy surrounding (a) the extent to which CARDS differs from standard ARDS, and (b) the resulting implications (if any) for clinical management. Case series and small observational studies early in the pandemic highlighted various atypical features of the disease [21–25], focusing in particular on the higher than usual compliance (and lower than usual amount of gassless tissue) for the same PaO_2_/FiO_2_ in early-stage CARDS patients. However, subsequent studies that compared larger cohorts of CARDS patients with “matched” cohorts of ARDS patients from previous studies failed to find significant differences [26,27]. A possible explanation for these discrepancies is that CARDS exhibits a dynamic time-dependent disease profile, encompassing several stages (or phenotypes), that starts with the highly atypical presentation described above and culminates in a more standard ARDS presentation as the disease progresses [28]. Thus, data from CARDS patients that was collected over different (or later, i.e., post-intubation) stages in the disease course could mask the particular characteristics of the early-stage presentation [29]. This argument is supported by a recent study of 114 exclusively early-stage (pre-intubation) CARDS patients that clearly showed the posited unique pathophysiological features of this disease, [30].

### Setting PEEP in invasive mechanical ventilation of CARDS patients

Several studies have examined lung recruitability and the effects of PEEP in CARDS patients. In a study of 12 patients with severe CARDS who had received various days of non-invasive or invasive ventilatory support before the first day of observation [42], predominantly (83%) poor recruitability was observed, as measured at the bedside using the recruitment to inflation ratio (R/I ratio) [43], and poorly recruitable patients were ventilated with PEEP values between 5 and 10 cmH_2_O. In [44], the authors performed multiple refined physiological measurements to evaluate recruitability in 10 CARDS patients at different time points along their clinical time course. Changing PEEP between 5 cmH_2_O and 15 cmH_2_O again revealed high inter-individual variability, with the increase in the lung volume due to a PEEP increase of 10 cmH_2_O varying from 16% to 140%. Despite this variability, driving pressure increased, and respiratory system compliance decreased, on average across the cohort in response to higher PEEP, indicating the potential for significant PEEP-induced overdistension in those patients with poorly recruitable lungs.

In [45], gas exchange, compliance and hemodynamics were assessed at two levels of PEEP (15 cmH_2_O and 5 cmH_2_O) within 36 h (day1) and from 4 to 6 days (day 5) after intubation in a cohort of 26 CARDS patients. These authors also found wide variation in recruitability, with PaO_2_/FiO_2_ ratio significantly increased at PEEP 15 cmH_2_O compared to 5 cmH_2_O only in the highly recruitable patients. Conversely, poorly recruitable patients exhibited a trend toward higher compliance at low PEEP compared to highly recruitable patients, again suggesting a risk of overinflation associated with high PEEP in these patients. The authors speculated that the small increase in PaO_2_/FiO_2_ ratio with PEEP observed in the poorly recruitable patients could be explained, at least in part, by a potential reduction in cardiac output induced by PEEP that may have contributed to decrease the shunt fraction.

A study, [46], of 17 patients with COVID-19 pneumonia fulfilling the Berlin criteria for severe ARDS on the 2^nd^ or 3^rd^ day of invasive mechanical ventilation evaluated respiratory mechanics, arterial blood gases, and hemodynamics before and after PEEP was reduced from settings based on standard ARDSnet criteria by an average of 29%. Reducing PEEP resulted in significantly increased respiratory system compliance and reduced hypercapnia.

PEEP reduction was not accompanied by lung derecruitment, and oxygenation did not deteriorate. The authors concluded that PEEP reduction decreased lung overdistension as interpreted by the increase in respiratory system compliance and decrease in dead space ventilation (reduced PaCO_2_). The authors note that, although a conservative or de-resuscitative fluid strategy is recommended in the management of patients with ARDS, in their CARDS patients application of PEEP levels based on the ARDSnet protocol was accompanied by substantial vasopressor dosage and 12-h fluid balance. PEEP de-escalation led to significant reduction of cumulative fluid balance during the following 12 hours and a three-fold decrease of vasopressor dosage. Decreased need for vasopressors and better fluid management translates into increased cardiac output and organ perfusion, accompanied by less fluid accumulation in the lungs.

In [3], respiratory mechanics were assessed in 14 CARDS patients, all of whom had higher than expected lung compliance compared to standard ARDS. Reducing PEEP increased lung compliance in all but one of the patients, and reduced dead space ventilation in all patients.

Finally, in [47], the effects of varying PEEP in 44 mechanically ventilated patients with severe COVID-19 pneumonia were analyzed via CT scan. Minimal alveolar recruitment was found to be induced by changes of PEEP from 8 cmH_2_O to 16 cmH_2_O. Higher PEEP improved oxygenation at FiO_2_ of 0.5 but not 1.0, and decreased respiratory system compliance.

### Setting PEEP in non-invasive pressure support ventilation of CARDS patients

The use of PEEP during non-invasive ventilation (NIV) of CARDS patients has been less well studied than in the case of invasive ventilation. In [48] inspiratory effort and respiratory mechanics in 30 spontaneously breathing patients receiving NIV for acute respiratory failure due to COVID-19 were compared with a “matched” cohort of standard ARDS patients. In both cohorts, mean PSV and PEEP values of 12 and 10 cm H_2_O, respectively, were reported. Application of NIV allowed CARDS patients to reduce their respiratory effort – measured as oesophageal pressure swings (ΔPes) from 12.4 to 7.6 cm H_2_O, and respiratory rate reduced from 28 to 24 breaths/min, after 2-hours of ventilatory support.

In a recent study of 114 early-stage CARDS patients treated with NIV, a recruitment manoeuvre involving increasing PEEP from 0 and 10 produced no improvement in oxygenation, but significantly increased an estimated value for total lung stress [30].

In [28], waveform traces are reported from an esophageal balloon catheter measuring esophageal pressure (Pes) as a surrogate of pleural pressure (Ppl) in two spontaneously breathing patients with SARS-CoV-2 pneumonia undergoing non-invasive ventilation (NIV) with Helmet. Computed tomography (CT) scans of both patients conformed to the proposed CARDS phenotype, i.e. ground glass opacities with limited amounts of alveolar collapse and atelectasis. Patient 1 presented with modest respiratory effort (ΔPes of 11 cm H_2_O, respiratory rate of 16 breaths/min) and subsequently exhibited progressive clinical and radiological improvement with ground glass resolutions, while Patient 2 had a significant respiratory effort (ΔPes of 32 cm H_2_O, respiratory rate of 35 breaths/min) and subsequently required intubation and invasive mechanical ventilation.

## Methods

### A computational simulator to compare CARDS versus ARDS pathophysiology

To shed further light on the above results, we employed a multicompartmental computational model that simulates highly integrated pulmonary and cardiovascular physiologies together with a detailed representation of the effects of mechanical ventilation. Computer simulation offers a fresh approach to traditional medical research that is particularly well suited to investigating treatment of critical illness. Critically-ill patients are routinely monitored in great detail, providing extensive high quality data-streams for model design & configuration and patient-matching. Models based on such data can incorporate very complex system dynamics that can be validated against patient responses for use as investigational surrogates. Crucially, simulation offers the potential to “look inside” the patient, allowing unimpeded access to all variables of interest. In contrast to trials on both animal models and human patients, *in silico* models are completely configurable and reproducible, for example, different ventilator settings can be applied to an identical virtual patient, or the same settings applied to different patients, in order to understand their mode of action and quantitatively compare their effectiveness.

Our simulator offers several advantages, including the capability to define hundreds of alveolar compartments (each with its own individual mechanical characteristics), with configurable alveolar collapse, alveolar stiffening, disruption of alveolar gas-exchange, pulmonary vasoconstriction and vasodilation, and airway obstruction. As a result, several defining clinical features of acute lung injury can be represented in the model, including varying degrees of ventilation perfusion mismatch, physiological shunt and deadspace, alveolar gas trapping with intrinsic PEEP, collapse-reopening of alveoli etc. The model has been successfully deployed in several previous studies investigating the pathophysiology and ventilatory management of conventional ARDS [31–36].

This model has recently been adapted to represent early-stage CARDS patients, [37]. Based on data [30] suggesting that early-stage COVID-19 patients have relatively well preserved lung gas volume and compliance, the model was set to have 8% of its alveolar compartments collapsed, i.e., non-aerated, by increasing the values of parameters representing alveolar extrinsic pressure and threshold opening pressure. To simulate the hyperperfusion of gasless tissue reported in [21,38,39], vasodilation was implemented in the collapsed units by decreasing their vascular resistance by 80%. Hypoxic pulmonary vasoconstriction (HPV) is normally incorporated in the simulator via a mathematical function - to simulate the hypothesised disruption of HPV in COVID-19, this function was disabled. Poor alveolar ventilation due to the effects of pneumonitis was modelled by disrupting alveolar–capillary gas equilibration in 20% of the alveolar compartments. As thrombotic complications have been reported to be a characteristic feature of COVID-19 [7,11,12,13,14], the presence of microthrombi was simulated by increasing vascular resistance by a factor of 5 in 10% of the compartments.

The cardiopulmonary simulator can be configured to represent either mechanically ventilated [37] or spontaneously breathing [40] CARDS patients.

### Simulating Mechanical Ventilation

Mechanical ventilation in the model is set to pressure-controlled mode with respiratory rate at 20 beats/min, inspiratory-to-expiratory ratio at 1:2, and Fio2 set to 100%. To observe the cardiopulmonary effects of interest, the following values were computed and recorded: *PaO*_2_, *PaCO*_2_, arterial pH, arterial oxygen saturation (*SaO*_2_), mixed venous oxygen saturation, volume of individual alveolar compartments at end-inspiration (*V*_*alv_insp*_), volume of individual alveolar compartments at end-expiration (*V*_*alv_exp*_), pressure of individual alveolar compartments at endinspiration (*P*_*alv_insp*_), pressure of individual alveolar compartments at end-expiration (*P*_*alv_exp*_), cardiac output (CO), mean arterial pressure, mean pulmonary artery pressure, and arterial oxygen delivery (*DO*_2_; computed using the values of CO, *SaO*_2_, and PaO_2_ and hemoglobin level). Alveolar recruitment was calculated as the fraction of alveoli receiving zero ventilation and subsequently achieving ventilation. VILI indices calculated include respiratory system compliance, intratidal recruitment, mechanical power [36], driving pressure, mean alveolar pressure, and dynamic strain (calculated as ΔV/V_frc_, where *V*_*frc*_ is *V*_*alv_exp*_ at PEEP = 0 *cm H*_2_*O* and Δ*V* = *V*_*alv_insp*_ − *V*_*alv_exp*_). Intratidal recruitment was calculated as the difference between the ratio of total ventilated lung at the end of inhalation to that at the end of exhalation. Shunt fraction (QS /QT) was calculated using the shunt equation, based on the arterial, pulmonary end-capillary, and mixed venous oxygen content. Physiologic dead space volume and tidal volume (*V*_*D*_/*V*_*T*_) were calculated from *PaCO*_2_, mixed expired *CO*_2_ tension, and exhaled *V*_*T*_ [41]. *PEEP*_*TOT*_ is measured as the total pressure within the lung at the end of expiration and accounts for both the extrinsic applied expiratory pressure and intrinsically generated end-expiratory pressures. Plateau pressure (*P*_*plat*_) is measured as the average end-inspiratory pressure in the lung. Lung compliance is calculated as ΔV/ΔP, where ΔP is the *P*_*plat*_ minus *PEEP*_*TOT*_. All simulations were run for 30 minutes, and during simulations, all variables were updated and recorded every 10ms.

### Simulating Non-Invasive Ventilation

The model was adjusted to allow for the simulation of non-invasive ventilation by running it in spontaneously breathing mode and then adding varying levels of pressure support from the ventilator. The simulator was configured to match the ΔPes waveforms from the two CARDS patients reported in [28], under spontaneous breathing with a standard NIV pressure support value of 12 cm H_2_O (Figure 1 panels (A) and (C)). We then examined the effect of applying varying PEEP levels while maintaining the spontaneous respiratory effort constant in each patient. The total lung stress reported is equal to the peak transpulmonary pressure as was reported by Coppola et al. [30].

**Figure 1:**
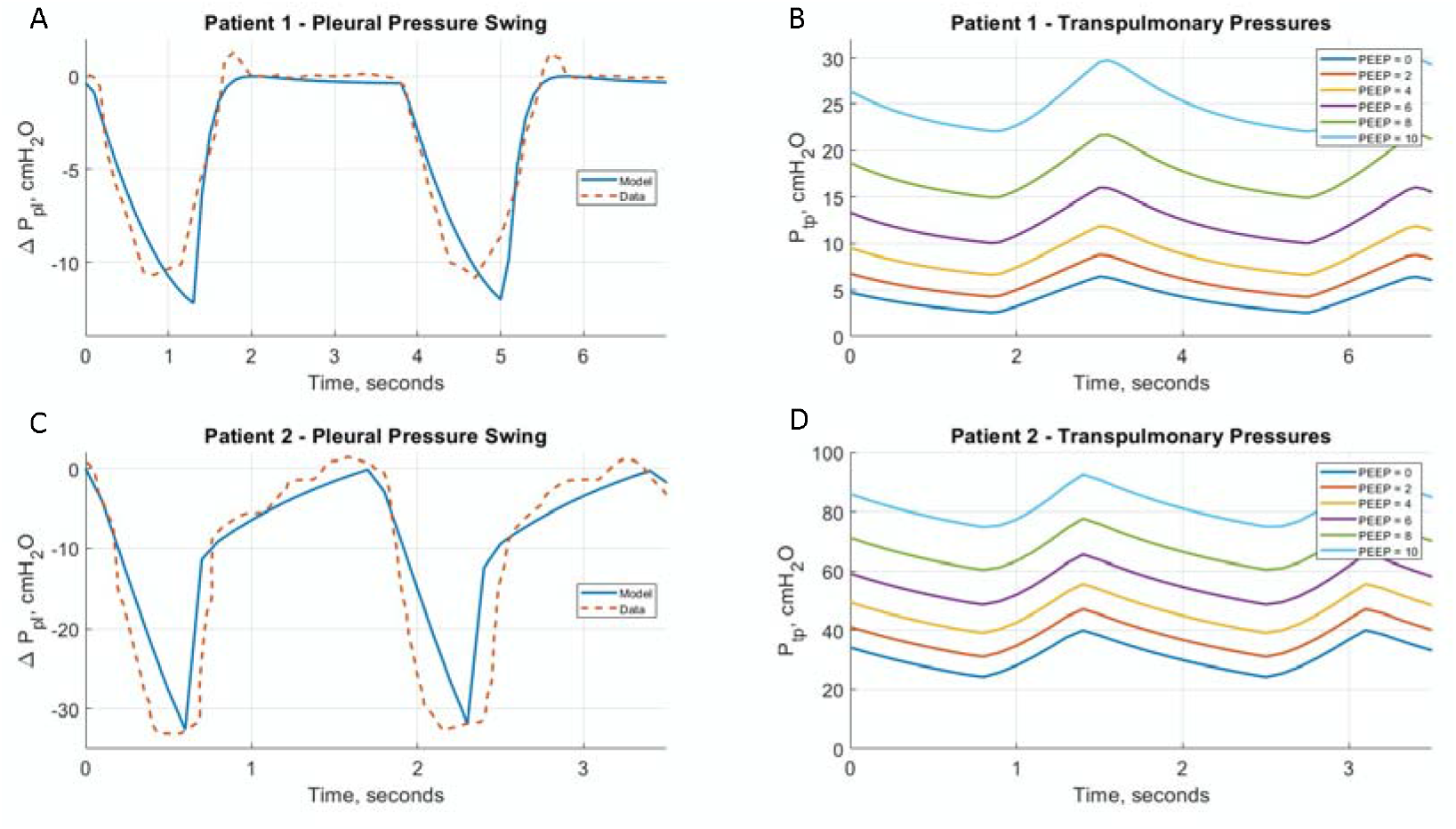
(A) Simulated pleural pressure swings from the CARDS model compared with ΔPes waveforms reported in [28], modest respiratory effort patient. (B) Simulated transpulmonary pressure swings for different levels of PEEP. (C) Simulated pleural pressure swings from the CARDS model compared with ΔPes waveforms reported in [28], high respiratory effort patient. (D) Simulated transpulmonary pressure swings for different levels of PEEP.

## Results

### Setting PEEP in invasive mechanical ventilation of CARDS patients

Although recent studies provide much compelling evidence, the interactions between haemodynamics (cardiac output and distribution of pulmonary blood flow) and alveolar recruitment with higher PEEP is difficult to establish at the bedside and can confound the discrepancy between changes seen in the oxygenation (improvement or deterioration) and the alteration in lung mechanics (either concordant or discordant). To shed further light on these issues, we configured our cardiopulmonary simulator to represent both the standard ARDS and hypothesised CARDS disease pathophysiology, as described in the previous section, and then compared its outputs, for the same ventilator settings, when PEEP is set at 5, 10 and 15 cm H_2_O. As shown in Figure 2, higher PEEP in the standard ARDS model leads to the typical benefits associated with recruitment of alveolar units in a lung suffering from significant levels of alveolar collapse – improved oxygenation, increased compliance, and reduced driving pressure. In the CARDS model, however, no oxygenation benefits are observable above a PEEP of 10 cm H_2_O, while compliance decreases and driving pressure increases as PEEP is increased. The change in lung volume ΔV (calculated as the end inspiratory lung volume - lung volume at FRC) is also significantly higher at each value of PEEP in the CARDS model compared to the standard ARDS model, indicating the potential for damaging levels of strain at higher PEEP.

**Figure 2:**
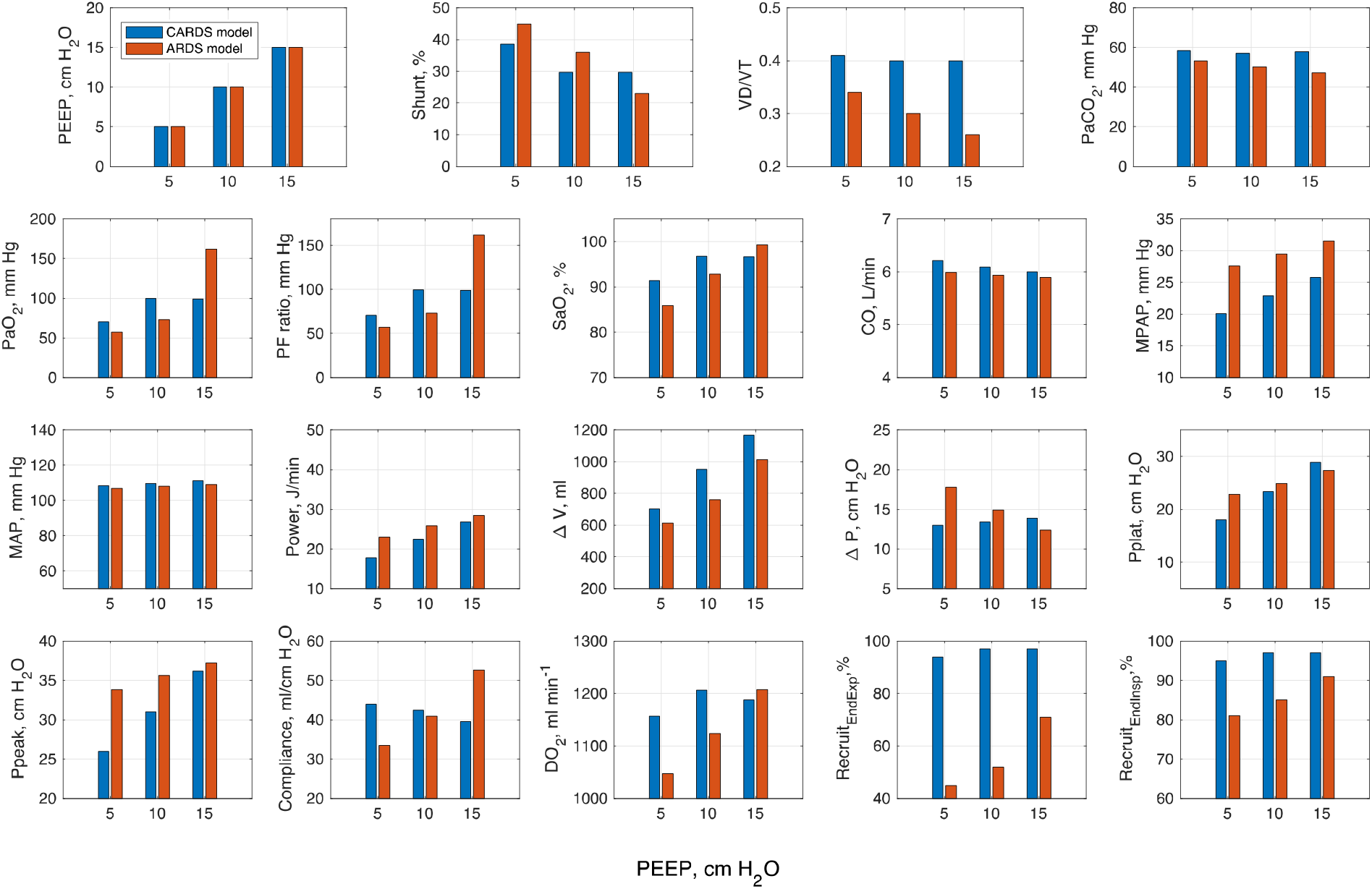
Simulated responses of CARDS and standard ARDS models to different levels of PEEP. Other mechanical ventilator settings were fixed at Tidal Volume = 470 ml (6.75 ml/Kg), Ventilation Rate = 20 breaths/min, Duty Cycle = 0.33, FiO2 = 100%.

### Setting PEEP in non-invasive pressure support ventilation of CARDS patients

As shown in Table 1, PEEP levels above 2 cm H_2_O produced no benefit in terms of oxygenation in either patient. The compliance of the lung and the respiratory system dropped markedly with increasing PEEP, while pleural and transpulmonary pressure swings (Figure 1 panels (B) and (D)), total stress and total strain all increased (Table 1). Interestingly, in [30], increasing PEEP from 0 and 10 also produced no improvement in oxygenation, while total lung stress was the only variable which was independently associated with negative outcome (intubation). In our simulations, an increase in PEEP from 0 to 10 produced large increases in total lung stress – from 6.5 to 29.8 cm H_2_O in the patient experiencing modest respiratory effort, and from 40.0 to 92.6 cm H_2_O in the patient experiencing significant respiratory effort (Table 1).

**Table 1:**
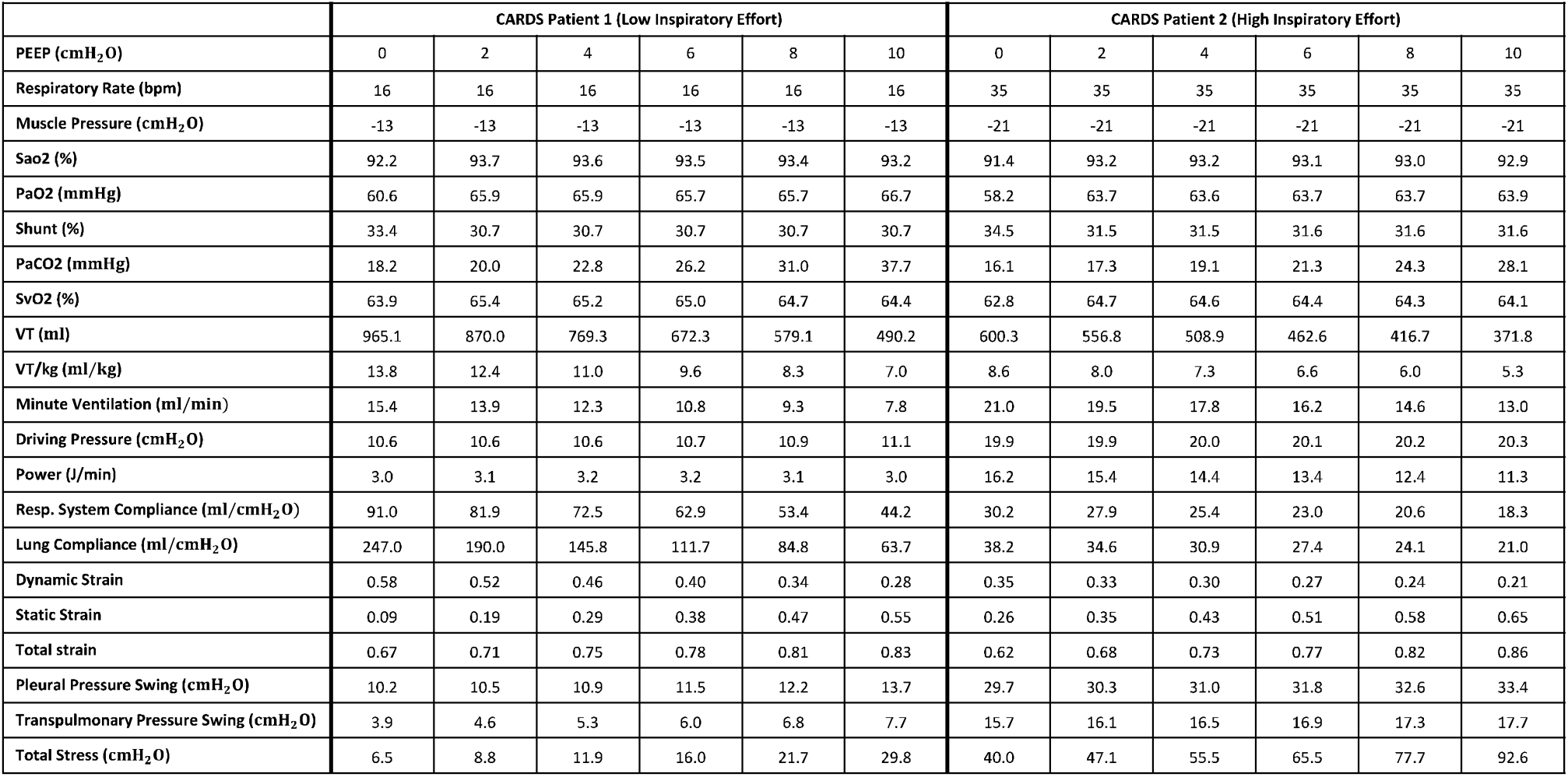
Simulated outputs for CARDS patients with moderate and high respiratory efforts undergoing NIV (based on data in [28]) with varying levels of PEEP.

|  | CARDS Patient 1 (Low Inspiratory Effort) |  |  |  |  |  | CARDS Patient 2 (High Inspiratory Effort) |  |  |  |  |  |
| --- | --- | --- | --- | --- | --- | --- | --- | --- | --- | --- | --- | --- |
| PEEP (cmH<sub>2</sub>O) | 0 | 2 | 4 | 6 | 8 | 10 | 0 | 2 | 4 | 6 | 8 | 10 |
| Respiratory Rate (bpm) | 16 | 16 | 16 | 16 | 16 | 16 | 35 | 35 | 35 | 35 | 35 | 35 |
| Muscle Pressure (cmH<sub>2</sub>O) | -13 | -13 | -13 | -13 | -13 | -13 | -21 | -21 | -21 | -21 | -21 | -21 |
| Sao<sub>2</sub> (%) | 92.2 | 93.7 | 93.6 | 93.5 | 93.4 | 93.2 | 91.4 | 93.2 | 93.2 | 93.1 | 93.0 | 92.9 |
| PaO<sub>2</sub> (mmHg) | 60.6 | 65.9 | 65.9 | 65.7 | 65.7 | 66.7 | 58.2 | 63.7 | 63.6 | 63.7 | 63.7 | 63.9 |
| Shunt (%) | 33.4 | 30.7 | 30.7 | 30.7 | 30.7 | 30.7 | 34.5 | 31.5 | 31.5 | 31.6 | 31.6 | 31.6 |
| PaCO<sub>2</sub> (mmHg) | 18.2 | 20.0 | 22.8 | 26.2 | 31.0 | 37.7 | 16.1 | 17.3 | 19.1 | 21.3 | 24.3 | 28.1 |
| SvO<sub>2</sub> (%) | 63.9 | 65.4 | 65.2 | 65.0 | 64.7 | 64.4 | 62.8 | 64.7 | 64.6 | 64.4 | 64.3 | 64.1 |
| VT (ml) | 965.1 | 870.0 | 769.3 | 672.3 | 579.1 | 490.2 | 600.3 | 556.8 | 508.9 | 462.6 | 416.7 | 371.8 |
| VT/kg (ml/kg) | 13.8 | 12.4 | 11.0 | 9.6 | 8.3 | 7.0 | 8.6 | 8.0 | 7.3 | 6.6 | 6.0 | 5.3 |
| Minute Ventilation (ml/min) | 15.4 | 13.9 | 12.3 | 10.8 | 9.3 | 7.8 | 21.0 | 19.5 | 17.8 | 16.2 | 14.6 | 13.0 |
| Driving Pressure (cmH<sub>2</sub>O) | 10.6 | 10.6 | 10.6 | 10.7 | 10.9 | 11.1 | 19.9 | 19.9 | 20.0 | 20.1 | 20.2 | 20.3 |
| Power (J/min) | 3.0 | 3.1 | 3.2 | 3.2 | 3.1 | 3.0 | 16.2 | 15.4 | 14.4 | 13.4 | 12.4 | 11.3 |
| Resp. System Compliance (ml/cmH<sub>2</sub>O) | 91.0 | 81.9 | 72.5 | 62.9 | 53.4 | 44.2 | 30.2 | 27.9 | 25.4 | 23.0 | 20.6 | 18.3 |
| Lung Compliance (ml/cmH<sub>2</sub>O) | 247.0 | 190.0 | 145.8 | 111.7 | 84.8 | 63.7 | 38.2 | 34.6 | 30.9 | 27.4 | 24.1 | 21.0 |
| Dynamic Strain | 0.58 | 0.52 | 0.46 | 0.40 | 0.34 | 0.28 | 0.35 | 0.33 | 0.30 | 0.27 | 0.24 | 0.21 |
| Static Strain | 0.09 | 0.19 | 0.29 | 0.38 | 0.47 | 0.55 | 0.26 | 0.35 | 0.43 | 0.51 | 0.58 | 0.65 |
| Total strain | 0.67 | 0.71 | 0.75 | 0.78 | 0.81 | 0.83 | 0.62 | 0.68 | 0.73 | 0.77 | 0.82 | 0.86 |
| Pleural Pressure Swing (cmH<sub>2</sub>O) | 10.2 | 10.5 | 10.9 | 11.5 | 12.2 | 13.7 | 29.7 | 30.3 | 31.0 | 31.8 | 32.6 | 33.4 |
| Transpulmonary Pressure Swing (cmH<sub>2</sub>O) | 3.9 | 4.6 | 5.3 | 6.0 | 6.8 | 7.7 | 15.7 | 16.1 | 16.5 | 16.9 | 17.3 | 17.7 |
| Total Stress (cmH<sub>2</sub>O) | 6.5 | 8.8 | 11.9 | 16.0 | 21.7 | 29.8 | 40.0 | 47.1 | 55.5 | 65.5 | 77.7 | 92.6 |

## Conclusions

The clinical studies performed to date, further confirmed by our simulations, indicate that use of standard protocols employing high PEEP in early-stage CARDS patients (whether invasively or non-invasively ventilated) is unlikely to lead to alveolar recruitment, and may be injurious in some patients due to the risk of significantly increasing transpulmonary pressure swings and total lung stress and strain. The limited improvement in oxygenation that can be observed in these circumstances can be conceptualised as the consequence of changes in haemodynamic (cardiac output) rather than alveolar recruitment per se.

In the context of CARDS patients undergoing NIV, the argument that high PEEP is necessary to allow patients to reduce their respiratory effort is not supported by the available data.

The emphasis on the atypical features of COVID-19 has been questioned by some authors, who point out that ARDS is by definition heterogeneous. It has also been shown that some of the alterations seen in CARDS are features that have already been encountered in ARDS from mixed aetiologies [49]. Although these general premises are certainly true and reasonable, the fact that the prevalence of these atypical features is much higher in COVID-19 than in ARDS should raise questions about the way that mechanical ventilation should be personalised, and what effects – beyond oxygenation – should be monitored. Specific pathophysiological considerations related to CARDS should also invite further clinical and physiological studies that evaluate the effects of PEEP on cardio-pulmonary interactions, recruitability, and inspiratory effort, as well as on how PEEP (or NIV) can alter the disease’s physiological trajectory over time. Ultimately, as we have learnt in the last 20 years of ARDS research, a strategy that leads to higher oxygenation may not be the most appropriate or safest strategy for every patient with ARDS, regardless of its aetiology.

## Data Availability

All data produced in the present work are contained in the manuscript

